# Lockdown exit strategies and risk of a second epidemic peak: a stochastic agent-based model of SARS-CoV-2 epidemic in France

**DOI:** 10.1101/2020.04.30.20086264

**Authors:** Nicolas Hoertel, Martin Blachier, Carlos Blanco, Mark Olfson, Marc Massetti, Marina Sánchez Rico, Frédéric Limosin, Henri Leleu

## Abstract

Most European countries have responded to the COVID-19 threat by nationwide implementation of barrier measures and lockdown. However, assuming that population immunity will build up through the epidemic, it is likely to rebound once these measures are relaxed, possibly leading to a second or multiple repeated lockdowns. In this report, we present results of epidemiological modelling that has helped inform policy making in France. We used a stochastic agent-based microsimulation model of the COVID-19 epidemic in France, and examined the potential impact of post-quarantine measures, including social distancing, mask-wearing, and shielding of the population the most vulnerable to severe COVID-19 infection, on the disease’s cumulative incidence and mortality, and on ICU-bed occupancy. The model calibrated well and variation of model parameter values had little impact on outcome estimates. While quarantine is effective in containing the viral spread, it would be unlikely to prevent a rebound of the epidemic once lifted, regardless of its duration. Both social distancing and mask-wearing, although effective in slowing the epidemic and in reducing mortality, would also be ineffective in ultimately preventing the overwhelming of ICUs and a second lockdown. However, these measures coupled with shielding of vulnerable people would be associated with better outcomes, including lower cumulative incidence, mortality, and maintaining an adequate number of ICU beds to prevent a second lockdown. Benefits would nonetheless be markedly reduced if these measures were not applied by most people or not maintained for a sufficiently long period, as herd immunity progressively establishes in the less vulnerable population.

## 1. Introduction

The COVID-19 pandemic is now a major global health threat. As of April 28, about 3 million confirmed cases and more than 200,000 deaths due to the novel severe acute respiratory syndrome coronavirus 2 (SARS-CoV-2) had been reported worldwide.^2^ Due to the lack of a vaccine or an effective treatment for COVID-19, most European countries have responded with a variety of non-pharmaceutical interventions (NPIs) intended to diminish the viral transmission by reducing contact rates in the general population.^3^ These measures include social distancing, wearing a face covering when outside of home, closing schools, churches, bars and other social venues, and all stores except groceries and pharmacies, screening of symptomatic people, and lockdown of suspected cases or of the full population. Countries in which these interventions have been implemented early in the epidemic have been successful at diminishing the number of incident cases and reducing the peak healthcare demand and deaths. However, assuming that population immunity will build up through the epidemic,^4^ it is likely to rebound once these measures are relaxed,^5–7^ as only a limited proportion of the European population will have been infected at this time,^8–10^ leading to the possibility of a second or even multiple repeated lockdowns. Such measures impose harmful burdens on the population and the global economy,^11^ and are difficult to tolerate during extended periods.^1^ Therefore, evaluating alternate NPIs that could be implemented at this stage and potentially avoiding a second epidemic peak and lockdown is urgently needed.^12,13^

To face the epidemic, France ordered on March 17 all nonessential retailers and services to close, and the general population to stay confined at home and to adhere to social distancing when outside of home for important personal needs. These measures have been successful in reducing the number of incident cases and the strain on the healthcare system, and the French government has announced a lockdown lifting on May 11th. However, in France as in many other countries where lockdown has been ordered, there is intense debate over which lockdown exit strategies should be implemented to avoid an epidemic rebound. Because COVID-19 is a newly emergent virus, for which much remains to be understood about its transmission and pathophysiology, model-based predictions of the public health impact of competing NPIs on the epidemic course are critical to help support evidence-based policy decisions.

In this report, we present results of epidemiological modelling that has helped inform policy making in France. Based on a stochastic agent-based microsimulation model^14^ of the COVID-19 epidemic in France, we have projected the potential impact of competing NPIs on the disease’s cumulative incidence and mortality, and on ICU-bed occupancy. Specifically, we evaluated quarantine extension from 8 to 16 weeks and post-quarantine measures including social distancing, mandatory mask-wearing, and shielding of the population the more vulnerable to severe COVID-19 infection. Advantages of ABM over other traditional modelling techniques include a flexible individual-based approach that can capture an emergent phenomenon with complex interactions between individuals in an heterogeneous population, and provide a natural description of a complex system.^14,15^ Because of several uncertainties that determine the risk of virus transmission, such as the number of asymptomatic cases and the duration of the contagious period,^12^ the present analysis followed recent recommendations for improving predictive mathematical models of the COVID-19 pandemic^16^ and was based on a calibration process that accounts for several disease’s transmission parameters within constraints defined by the contact matrix and known parameters of the disease.

## 2. Methods

Following previously described methods,^14^ which have been recently applied to model the COVID-19 epidemic in New York City,^17^ we built a stochastic ABM model of the epidemic of COVID-19 in France. The model included 194 parameters summarized in **eTable 1**. Parameters on individual and disease characteristics (n=161) were mainly based on available data from prior studies and model calibration. Parameters related to social contacts were based on either prior studies (n=11) or assumptions when no data were available (n=22). The source code of the model has been deposited in a recognized public source code repository (GitHub).

### 2.1. Individuals’ characteristics

The model was built to reproduce the household (proportions of singles, couples with children, couples without children, and single parents with children) and age structures (categorized by 5-year age groups) observed in the French general population.^18^ Households were distributed on a square grid that represents a geographical area approximating France. Based on age- and sex-stratified national estimates,^19^ all subjects were attributed a probability of having one or multiple conditions known to increase the risk of severe SARS-CoV-2 infection,^8,20,21^ including obesity, hypertension, diabetes, coronary diseases, and chronic pulmonary diseases (**eTable 1**). Individuals with a least one of these conditions or aged over 65 years were considered to be part of the population the most vulnerable to severe COVID-19 infection.^8,20,21^ Based on the age distribution and national estimates of these conditions in France,^22^ we calculated that this population represents more than 30% of the French general population.

### 2.2. Social contacts

Social contacts were modelled to enable specific restrictions due to quarantine (e.g., school closure, cancellation of public events), while conserving unavoidable contacts such as with intrafamilial members or grocery shopping during the quarantine period. Given the complexity of modelling social contacts, we used a simplified set of contacts at both individual and household levels to model different types of social contacts experienced during the day.^23^ These included close contacts for a prolonged duration with a small number of individuals, such as intrafamilial contacts, or people met at school or work. They also included less frequent and less prolonged contacts with a finite set of individuals such as friends or extended family members. Finally, they included brief contacts with individuals in centralized locations such as grocery shopping, or in more remote locations such as when using public transport. For details on the parameter values used in the model to reproduce social contacts, please refer to *Supplemental text section*.

### 2.3. SARS-CoV-2 characteristics

SARS-CoV-2 characteristics were based on reports from Santé Publique France,^19^ Institut Pasteur,^24^ the European Centre for Disease Prevention and Control (ECDC),^25^ and the London Imperial College.^26^ For details on the parameter values used in the model, please refer to *Supplemental text section*.

### 2.4. Medical outcomes

Medical outcomes included cumulative incidence, cumulative mortality, and number of ICU beds needed.

### 2.5. Interventions

All diagnosed cases were assumed to be quarantined. In the model, we also took into account efforts to track the contacts of diagnosed patients. Every intrafamilial, friend and family, work, and school contact of a diagnosed patient had in the previous days was considered to be systematically tested with RT-PCR after an average delay of two days, representing the delay of the investigation. During this period, infected contacts could further spread the infection. People who met in grocery stores or in public transports were assumed to be untraceable. During the lockdown, we considered that individuals had no contacts with other people, except with intrafamilial members and individuals at random in grocery stores and in streets. Finally, based on prior work,^27,28^ it was assumed that the risk of transmission between individuals would be decreased by 50% if all individuals either adhered to social distancing or were wearing masks, and that both measures would reduce this risk multiplicatively by 75%.

Based on opening statements from the French government, we considered in all scenarios that (i) the quarantine and restrictions for school, work and public transport will be lifted on May 11^th^, (ii) restaurants and bars will remain closed from May 11^th^ until June 11^th^, and (iii) attendance to cinemas, museums and public events will be authorized on July 11^th^.

We successively examined the following scenarios, using a « stepped care » approach:

- i) The natural course of the epidemic if no quarantine had been ordered.
- ii) Two different durations of quarantine: 8 weeks, i.e. the quarantine period scheduled in France, and an 8-week extension, i.e. 16-week quarantine.
- iii) Post-quarantine protection measures for all individuals, including social distancing and mask-wearing.
- iv) Post-quarantine shielding of individuals vulnerable to severe SARS-CoV-2 infection, i.e., individuals aged over 65 years or having comorbidity, including obesity, hypertension, diabetes, coronary diseases, or chronic pulmonary diseases. Shielding implied that individuals always stayed home except for grocery shopping, could stay with family members living in their home but with protection measures (i.e., social distancing and masks), did not attend any gatherings, including gatherings of friends and families living outside, and strictly avoided contact with people displaying symptoms of COVID-19. We also evaluated the effect of both the duration of this intervention and the proportion of vulnerable people shielded on medical outcomes.

### 2.6. Statistical analyses

The stochastic agent-based microsimulation model (ABM) was run from March 1, 2020 until the end of December, 2020, on 500,000 individuals. It was performed on April, 11^th^ using data obtained until April 10^th^. The results were extrapolated to the French population, which comprises 67 million people. We examined whether the model had adequate calibration based on both two-sample Kolmogorov–Smirnov (KS) tests and visual comparison of the model-predicted and observed curves of the cumulative incidence of ICU admissions, ICU-bed occupancy, and cumulative mortality. Because the model included 500,000 individuals approximating 67 million people from the general population, the maximum possible precision was ±134 individuals. Therefore, KS tests were performed after correcting the observed data with the available precision, i.e. dividing and rounding the modeled and observed data by the precision. We also compared the model-predicted and observed age distribution among deceased people using both a Cochran-Mantel-Haenszel test and visual comparison. Finally, we examined whether the model-predicted value for the R_0_ of COVID-19 at the onset of the quarantine in France (i.e., March 17^th^) was in line with published reports.

Following recent recommendations for improving predictive mathematical models of the COVID-19 pandemic,^16^ we examined the robustness of our results by evaluating the impact on the estimated cumulative incidence and mortality, and the number of ICU beds needed of varying each model parameter value by +/-20%. These analyses were run for the combination of post-quarantine social distancing and mask-wearing for the general population, and shielding of vulnerable individuals.

The model was performed using C++ and statistical analyses were conducted using SAS 9.4. The threshold for statistical significance was a priori fixed at p<0.05.

## 3. Results

### 3.1. Model calibration

**Figure 1** presents the results of the model calibration, supporting visually a good fit between observed and model-predicted cumulative incidence of ICU admissions, ICU-bed occupancy, cumulative mortality, and age distribution of deceased people. After correcting for precision, two-sample Kolmogorov–Smirnov tests comparing observed and model-predicted curves of the cumulative incidence of ICU admissions, ICU-bed occupancy, and the cumulative mortality did not show significant differences [KSa=0.12 (p=0.99), KSa=0.23 (p=0.99), and KSa=1.01 (p=0.25), respectively], as did the Cochran-Mantel-Haenszel test comparing observed and model-predicted age distribution of deceased people (χ^2CMH^=0.34, p=0.55). Finally, the R_0_ of COVID-19 predicted by our model was 3.1 at the onset of the quarantine, consistent with the findings of a review^29^ suggesting that R_0_ estimates would range between 1.40 and 6.49, with a median of 2.79.

**Figure 1.**
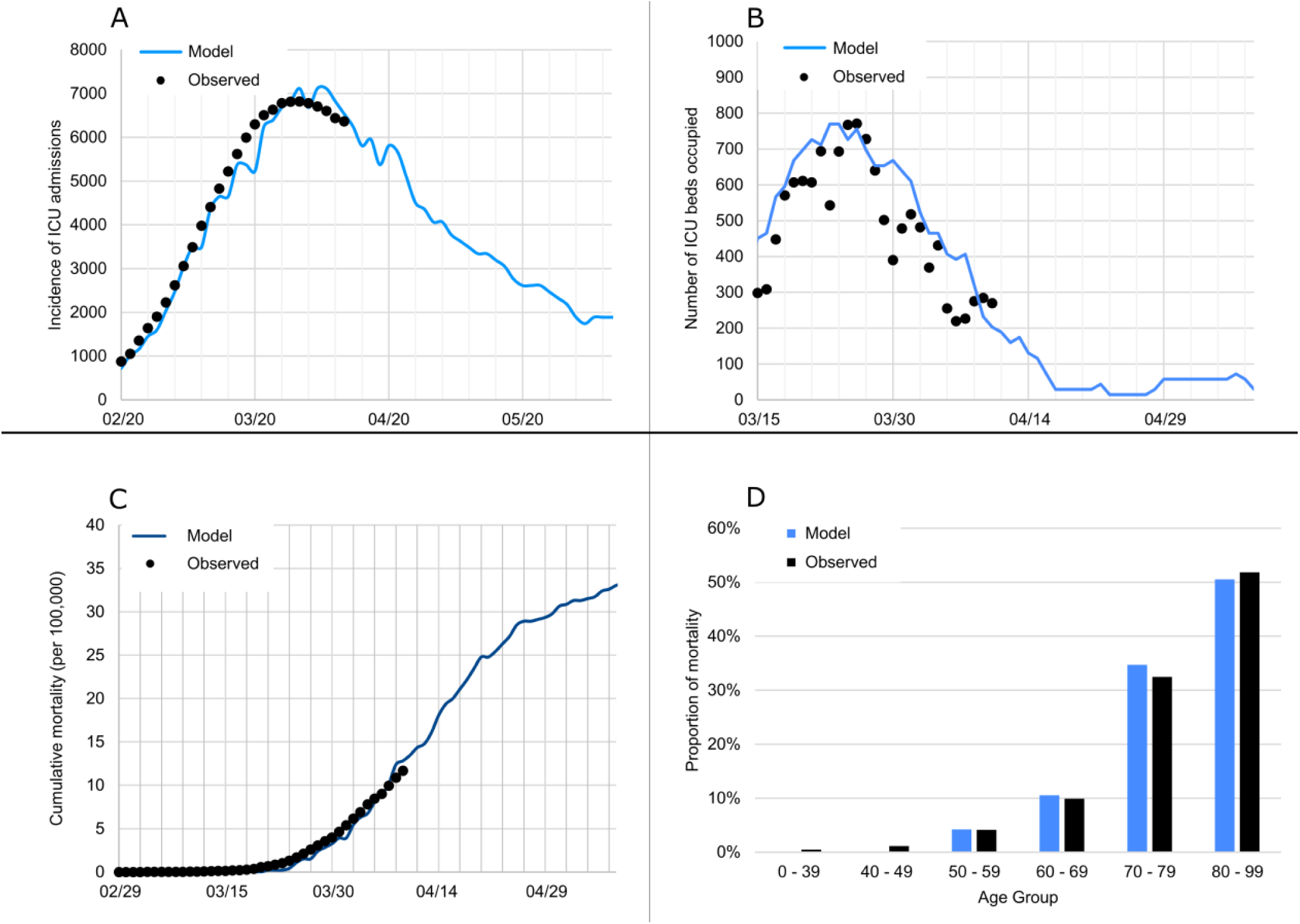
Model-predicted and observed curves of the cumulative incidence of ICU admissions (A), ICU-bed occupancy (B), cumulative mortality (C), and age distribution of deceased people (D) in France.

### 3.2. Effect of quarantine duration

While quarantine is highly effective at containing viral spread, we projected that it would be unlikely to prevent a second epidemic peak once lifted, regardless of its duration. Based on our model, the duration of quarantine (i.e., 8 or 16 weeks) alone was not associated with a reduced cumulative COVID-19 incidence or mortality, resulting in a similar, albeit delayed, overwhelming of ICUs, likely to lead to a second lockdown (**Figure 2; Table 1**).

**Figure 2.**
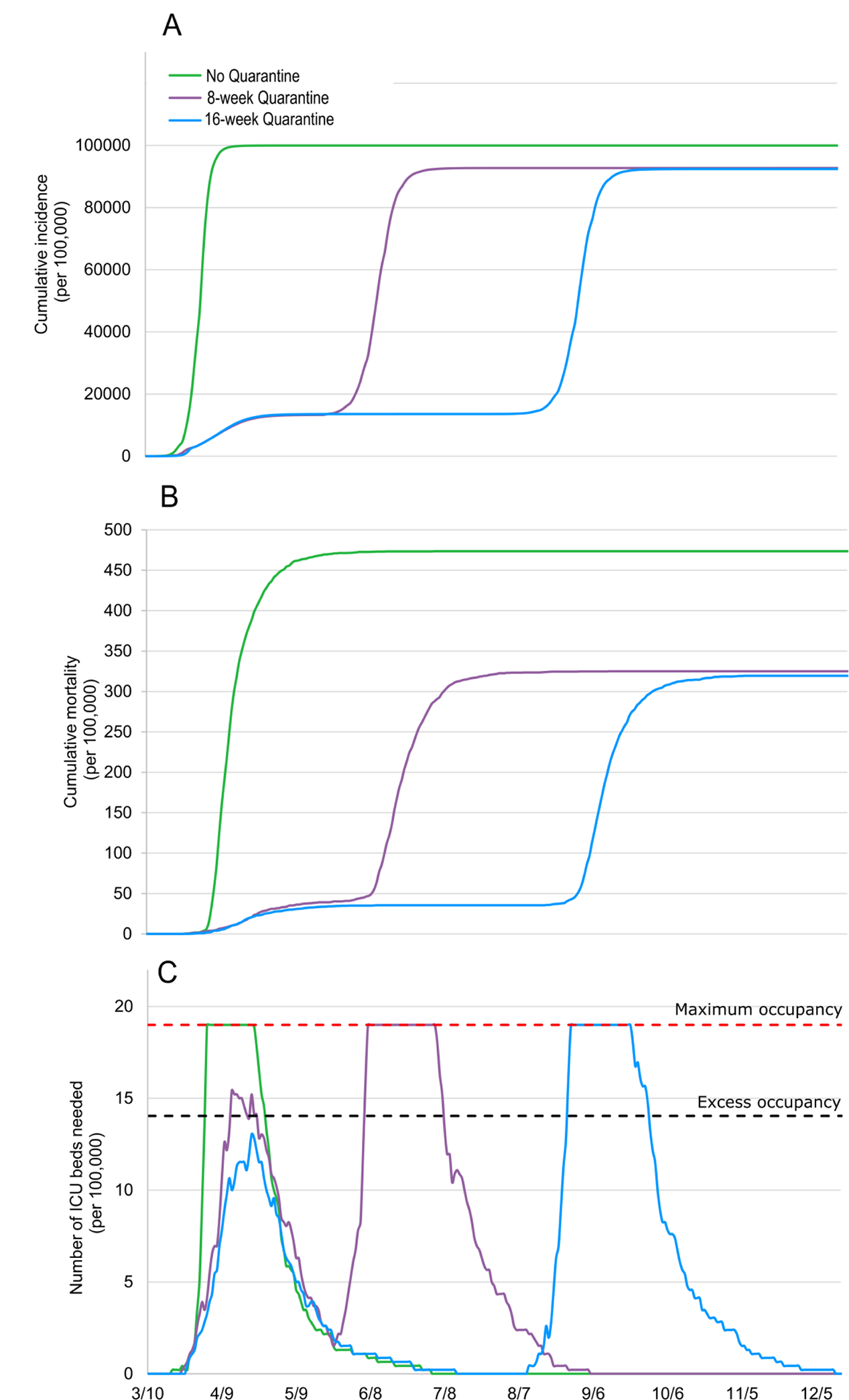
Model-predicted cumulative incidence (A), mortality (B), and number of ICU beds needed (C) by quarantine duration.

**Table 1.** Summary of projected effects of interventions on the COVID-19 cumulative incidence and mortality at the end of December 2020.

| ***Intervention*** | **Cumulative incidence** |  | **Cumulative mortality** |  |  |
| --- | --- | --- | --- | --- | --- |
|  | Absolute number<br>per 100,000 people | Percentage<br>reduction | Absolute number<br>per 100,000 people | Absolute number<br>in France | Percentage<br>reduction |
| 8-week quarantine + no specific post-quarantine measures | 93,000 | *Reference* | 325 | 217,750 | *Reference* |
| 16-week quarantine + no specific post-quarantine measures | 93,000 | 0% | 320 | 214,400 | 2% |
| 8-week quarantine + social distancing | 85,000 | 9% | 260 | 174,200 | 20% |
| 8-week quarantine + social distancing + mask-wearing | 65,000 | 30% | 130 | 87,100 | 60% |
| 8-week quarantine + social distancing + mask-wearing +<br>16-week shielding of 100% of vulnerable people | 60,000 | 35% | 115 | 77,050 | 65% |
| 8-week quarantine + social distancing + mask-wearing +<br>38-week shielding of 50% of vulnerable people | 60,000 | 35% | 105 | 70,350 | 68% |
| 8-week quarantine + social distancing + mask-wearing +<br>38-week shielding of 75% of vulnerable people | 54,000 | 42% | 70 | 46,900 | 78% |
| 8-week quarantine + social distancing + mask-wearing +<br>38-week shielding of 100% of vulnerable people | 40,000 | 57% | 50 | 33,500 | 85% |
|  | Absolute number<br>per 100,000 people | Percentage<br>augmentation | Absolute number<br>per 100,000 people | Absolute number<br>in France | Percentage<br>augmentation |
| 8-week quarantine + no specific post-quarantine measures | 93,000 | *Reference* | 325 | 217,750 | *Reference* |
| No quarantine | 100,000 | 7% | 475 | 318,250 | 32% |

### 3.3. Effect of post-quarantine social distancing and mask-wearing

We found that maintaining social distancing after ending the quarantine would be associated with a substantial slowdown of the epidemic, as shown by a flattening of the cumulative incidence curve, and a 20% decrease in cumulative mortality, after 8-week quarantine (**Figure 3; Table 1**). Combining social distancing and mandatory mask-wearing further flattened the epidemic curve and would be associated with an additional 40% decrease in mortality, corresponding to a 60% reduction of mortality compared to the absence of post-quarantine protection measures. However, although effective in slowing the epidemic and in reducing mortality, we found that this combination of measures would also be ineffective to prevent a second epidemic peak, likely to exceed ICU bed capacity and to lead to a second lockdown.

**Figure 3.**
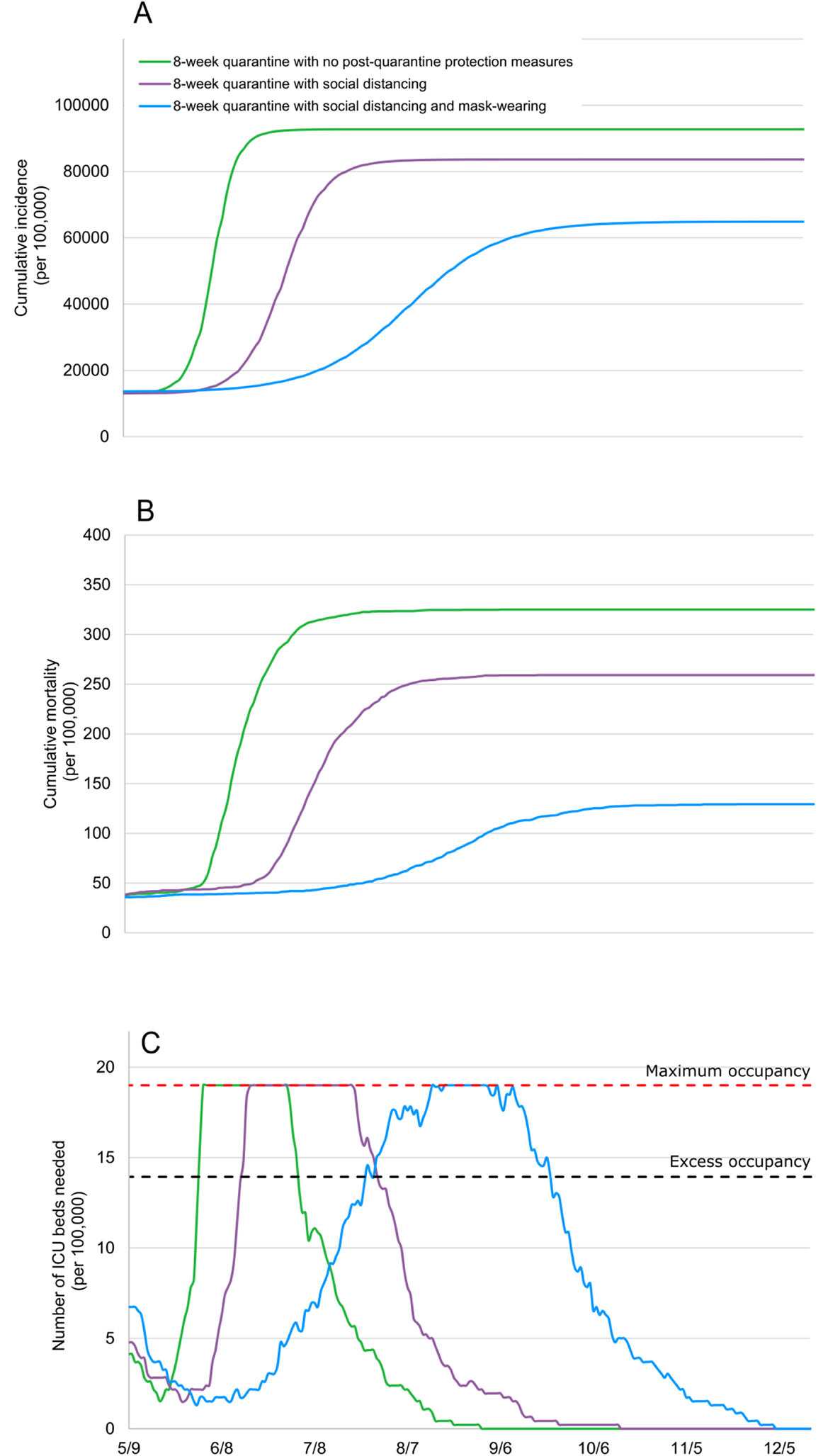
Model-predicted cumulative incidence (A), cumulative mortality (B), and number of ICU beds needed (C) associated with post-quarantine social distancing and mask-wearing for the general population.

### 3.4. Effect of post-quarantine shielding of people the most vulnerable to severe SARS-CoV-2 infection

We projected that the shielding of vulnerable people until the end of the epidemic (estimated in our model at 38 weeks after the quarantine lifting with this scenario) in addition to post-quarantine social distancing and mask-wearing for all individuals would be associated with a substantial slowdown of the epidemic, as shown by a strong flattening of the cumulative incidence curve and a substantial decrease in mortality of 62% compared with post-quarantine social distancing and mask-wearing only, and of 85% compared with the absence of specific post-quarantine intervention (**Figure 4; Table 1**).

**Figure 4.**
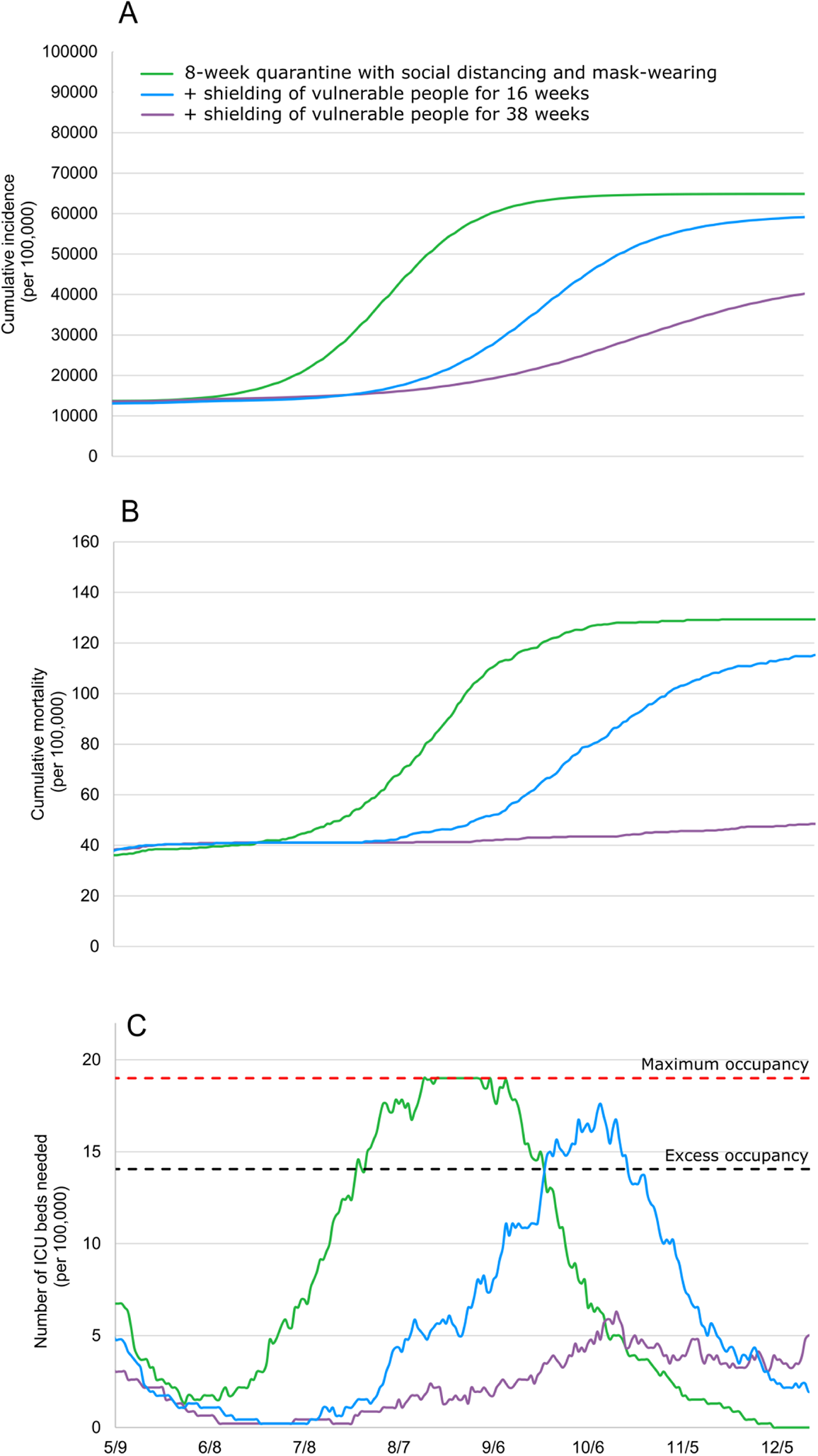
Model-predicted cumulative incidence (A), cumulative mortality (B), and number of ICU beds needed (C) associated with post-quarantine shielding of vulnerable individuals.

Furthermore, combining these 3 interventions would prevent an overwhelming of ICU capacity and substantially reduce mortality, but only if the interventions are maintained for a sufficiently long period and applied by most people. For example, interrupting the shielding of vulnerable people 16 weeks after the quarantine lifting would result in an increased risk of ICU overwhelming and a decrease in mortality of 12% compared with post-quarantine social distancing and mask-wearing only, and of 65% compared with the absence of specific post-quarantine intervention (**Figure 4; Table 1**). Similarly, partial adherence to shielding, defined as having only 50% of the vulnerable population shielded, would lead to a decrease in mortality of 19% compared with post-quarantine social distancing and mask-wearing only, and of 68% compared with the absence of specific post-quarantine intervention, and would not be sufficient to prevent a second epidemic peak, likely to lead to a second lockdown (**Figure 5; Table 1**).

**Figure 5.**
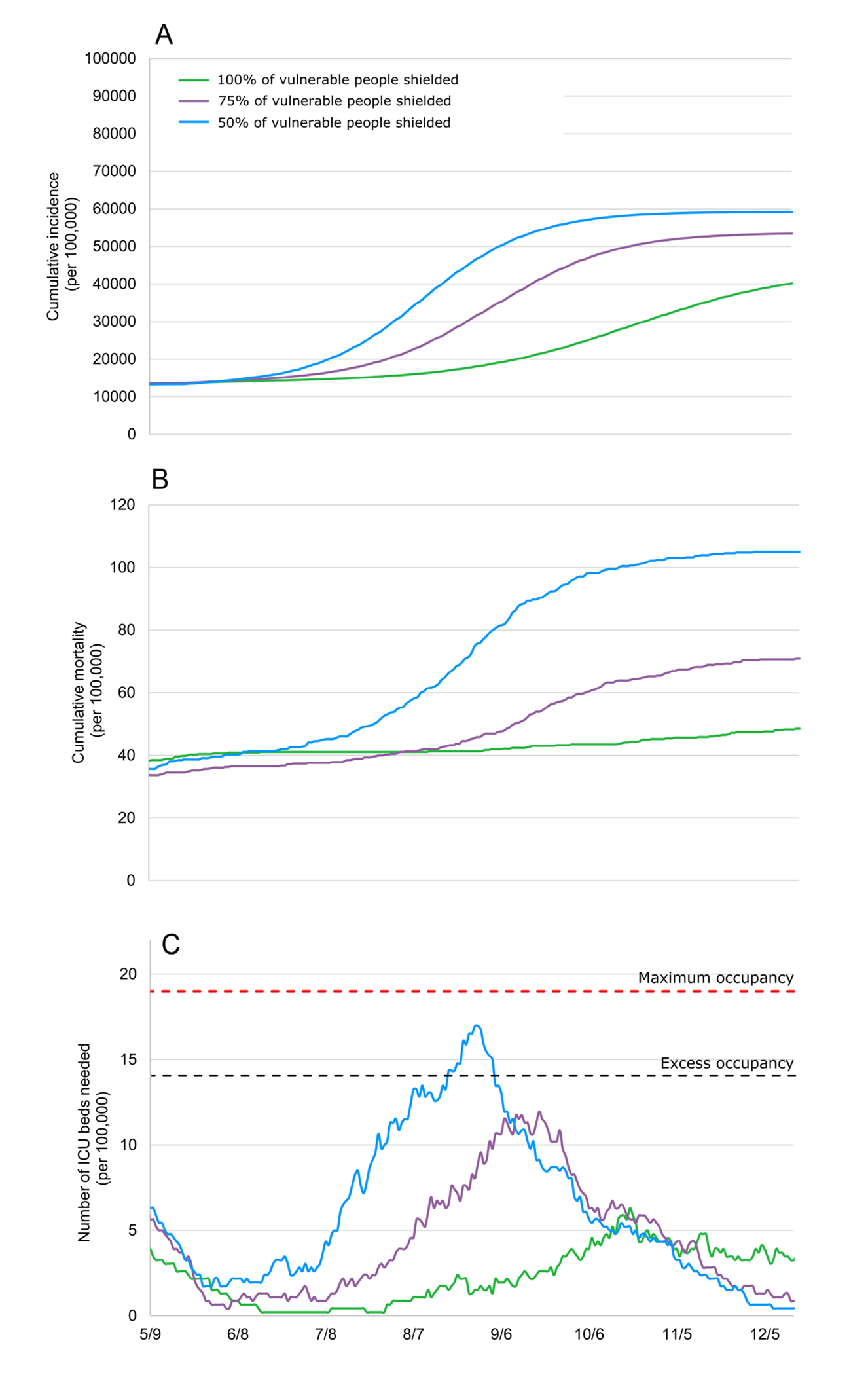
Model-predicted cumulative incidence (A), cumulative mortality (B), and number of ICU beds needed (C) according to the proportion of vulnerable individuals shielded.

### 3.5. Sensitivity analyses

By varying each model parameter value by +/-20% for the scenario combining post-quarantine social distancing, mask-wearing, and shielding of vulnerable individuals, we found that it would change COVID-19 cumulative incidence by at most 2,200 per 100,000, mortality by 42 per 100,000, and ICU-bed occupancy by 12 per 100,000, suggesting the robustness of our results (**eFigures 1 to 3**).

## 4. Discussion

To our knowledge, this is the first study to propose an agent-based microsimulation model of the epidemic of COVID-19 in France to predict the potential impact of post-quarantine measures, including social distancing, mask-wearing, and shielding of the population the most vulnerable to severe COVID-19 infection on the disease’s cumulative incidence, mortality, and on ICU-bed occupancy. The model calibrated well and the variation of each model parameter value by ±20% had limited impact on outcome estimates, suggesting the robustness of our results. While quarantine is a highly effective means of containing viral spread, it would be unlikely to prevent a rebound of the epidemic and the need for a second lockdown once lifted, regardless of its duration. Social distancing and mask-wearing, when implemented in association, would be very effective in slowing down the epidemic and in reducing mortality, but would be ineffective to ultimately prevent an overwhelming of ICU bed capacity. However, these measures when coupled with shielding of people the most vulnerable to severe SARS-CoV-2 infection, would be associated with better outcomes than these measures without shielding, including a lower cumulative incidence, mortality and number of ICU beds needed, sufficiently to prevent an overwhelming of ICU bed capacity and a second lockdown. Benefits would nonetheless be substantially reduced if these measures were not applied by most people or not maintained for a sufficiently long period, as herd immunity progressively establishes in the less vulnerable population.

Our findings reinforce that SARS-CoV-2 infection represents a major public health threat in France. This disease may cause a very high number of deaths, estimated by our model at more than 300,000 deaths in France alone if no quarantine had been ordered and if ICUs had been overwhelmed. It further predicts more than 200,000 deaths if no specific mitigation measures are planned at the time of quarantine lifting. In line with prior work^5–7^, our findings suggest that quarantine, while a highly effective strategy to reduce the strain on healthcare systems by delaying the epidemic peak, is unlikely, if applied as a standalone strategy, to prevent the rebound of the epidemic and the need of a second lockdown. In line with this prediction, we found that combining post-quarantine social distancing and mask-wearing, while quite effective in reducing mortality by decreasing ICU-bed saturation and the R_0_ of the COVID, would be ineffective in ultimately preventing ICU bed capacity from becoming overwhelmed and a subsequent second lockdown.

By contrast, we found that coupling these measures with shielding people the most vulnerable to severe SARS-CoV-2 infection would be associated with better outcomes than only implementing social distancing and wearing of masks. These improved outcomes include a lower mortality and number of ICU beds needed, in a sufficient magnitude to prevent overwhelming ICU bed capacity and a second lockdown. Specifically, mortality would be reduced up to 85% if this scenario is applied by 100% of vulnerable people until the end of the epidemic compared to the absence of specific measures after lifting the quarantine. Benefits would nonetheless be dramatically reduced if these measures were not maintained for a sufficiently long period and not applied by most vulnerable individuals, because it would result in an lesser decrease in mortality and increased risk of ICU overwhelming and of the need of a second lockdown.

Shielding vulnerable individuals implied in our simulation that these individuals always stayed home, except for grocery shopping, could stay with family members living in their home but with protection measures (i.e., social distancing and masks), did not attend any gatherings of friends and family members living outside their home, and strictly avoided contact with people presenting symptoms of COVID-19. Because prolonged quarantine exposes to increased risk of psychological and medical complications,^1^ we consider that this less stringent strategy, while ensuring an adequate protection of this vulnerable population whose proportion is estimated at more than 3 of 10 people in France, would have better chance of being applied by most individuals. However, it is crucial to provide clear rationale and information for these measures, appeal to altruism by reminding people of the benefits to wider society, and ensure sufficient supplies and adequate healthcare access are provided.

Our results could be explained by the herd immunity effect. It corresponds to the reduction of infection rates as a result of the indirect protection observed in the unimmunized segment of the population in which a large proportion has been infected and therefore immunized.^4^ This is reflected by the predicted flattening of the cumulative incidence below 70% of the population with this strategy. Indeed, by slowing down to a far greater extent the viral spread in the vulnerable population than in the healthiest population, most of infected people would likely be individuals who are at lower risk to developing severe or critical symptoms^5^ and being adequately treated since ICU capacity is not expected to be overwhelmed, even during the peak incidence. Once a majority of the low-risk population have been immunized against COVID-19, the herd immunity effect would be likely to prevent vulnerable people from becoming infected. Because protection measures would be used by the whole population to flatten the epidemic rebound, however, the herd immunity effect, possibly associated with the progressive extinction of the epidemic, would be reached only at 38 weeks after the quarantine lifting, i.e., the end of February, 2021.

Our study has several limitations. First, the model was calibrated on the diagnosis and mortality rates available from Santé Publique France^19^ and Institut Pasteur.^24^ However, we cannot exclude the possibility that these parameters are biased, as asymptomatic undiagnosed patients are likely responsible for a large hidden epidemic. Nevertheless, the observed differences across scenarios remained unchanged when considering a much higher and unlikely^8–10^ diagnosis rate of 1 in 10, supporting the robustness of our conclusions. Second, the contact matrix was approximated using multiple assumptions for each type of contact. However, we found that the model calibrated well, suggesting that it might adequately predict the course of the COVID-19 epidemic in France. Third, following standard assumptions, we considered that infected people could develop immunity for at least several months. However, post-COVID-19 immunity length remains unknown. Fifth, the impact of many of mitigation measures depends on how people react and adhere to them, which is likely to vary across segments of the populations. Finally, as with any simulation model, the results should be interpreted as estimates.

SARS-CoV-2 represents a major public health threat in France and worldwide. Post-quarantine social distancing and wearing of masks for the whole population, coupled with shielding of vulnerable people would substantially lower mortality and prevent a second lockdown. If these measures are applied by most people or maintained for a sufficiently long period, they will provide time for herd immunity to become progressively established in the less vulnerable population.

## Data Availability

The source code of the model has been deposited in a recognized public source code repository (GitHub).

## Acknowledgments

We thank Pr Melanie Wall and Pr Yuanjia Wang for their helpful comments on early versions of the methods.

## SUPPLEMENTRY METERIAL

**eTable 1.** Summary of main model parameters.

| Parameters | Value | Source |
| --- | --- | --- |
| **Individuals' characteristics** |  |  |
| Family structure (%) |  | Insee, 2020 <sup>1</sup> |
| Singles | 37% |  |
| Couples with children | 27% |  |
| Couples without children | 27% |  |
| Singles with children | 9% |  |
| Age structure (categorized by 5-year age groups) |  | Insee, 2020 <sup>1</sup> |
| Condition or disease associated with increased risk of death from SARS-COV2 (i.e., hypertension, diabetes, obesity, heart diseases, chronic pulmonary disease, and end-stage renal disease) | Estimates per 10-year age groups and sex | 2-7 |
| **Social contacts** |  |  |
| School class size (average) | 30 | Assumption |
| Proportion of small companies (<10 employees) | 18.40% | INSEE, 2018 <sup>8</sup> |
| Number of colleagues in small companies (average) | 2 | Assumption |
| Number of colleagues in bigger companies (average) | 10 | Assumption |
| Employment rate (for people aged 20 to 65 years) | 92% | INSEE, 2020 <sup>8</sup> |
| Shopping density (per 100,000 inhabitants) | 29.3 | APUR, 2018 <sup>9</sup> |
| Number of shopping trips (average per week) | 1.2 | Assumption |
| Number of people met per shopping trip (average) | 5 | Assumption |
| Social network distance | 22 | <sup>10</sup> |
| Frequency of meeting friends (average per week) | 1 | Assumption |
| Event participations, i.e., museum, cinema, music and sport events (average per year) | 5.4 | French Ministry of Culture, 2015 <sup>11</sup> |
| Close encounters per event participation (average) | 5 | Assumption |
| Round trips with public transport (average per week) for workers | 5 | Assumption |
| Round trips with public transport (average per week) for non-workers | 1.7 | Assumption |
| Close encounters in public transport | 3-5 | Assumption, with work-related trips assumed to happen at peak times with more encounters |
| International contamination (average, per week) | 1.8 | Based on Imported Case Frequency observed in France initially <sup>7</sup> |
| **SARS-CoV-2 infection characteristics** |  |  |
| Contamination risk (per min/m<sup>2</sup>) | 0.028 | Calibrated on the observed cumulative incidence of confirmed cases in France based on reports from Santé Publique France <sup>7</sup> and Institut Pasteur <sup>12</sup> |
| Proportion of asymptomatic/paucisymptomatic infections that will not be diagnosed | [100%-98%] | Assumption |
| Proportion of asymptomatic cases among people with asymptomatic or paucisymptomatic infection | 25% | London Imperial College, 2020 <sup>13</sup> |
| Hospitalization rates | [0.1%-31.4%] | Institut Pasteur <sup>12</sup> |
| ICU rates (if hospitalized) | [3.4%-36.4%] | Institut Pasteur <sup>12</sup> |
| Mortality rates (if hospitalized) | [0%-42%] | Institut Pasteur <sup>12</sup> |
| HR of obesity for death | 1.56 [1.32-1.84] | <sup>14</sup> |
| HR of diabetes for death | 1.29 [1.09-1.53] | <sup>14</sup> |
| Interaction diabetes*age<40 | 3.13 [1.68-5.81] | <sup>14</sup> |
| HR of COPD for death (COPD) | 1.50 [1.14-1.96] | <sup>14</sup> |
| HR of hypertension for death | 1.40 [1.17-1.63] | <sup>14</sup> |
| Delays (days) |  |  |
| Incubation time (average, standard deviation) | 6.4 (2.3) | <sup>15</sup> |
| Infection onset to diagnosis (average, standard deviation) | 2.1 (2.6) | <sup>16</sup> |
| Infection onset to hospital admission (average, standard deviation) | 5.8 (4.2) | London Imperial College <sup>13</sup> |
| Infection onset to recovery (average, standard deviation) | 20.5 (6.7) | London Imperial College <sup>13</sup> |
| Infection onset to death (average, standard deviation) | 16.0 (8.21) | London Imperial College <sup>13</sup> |
| Hospital to ICU (average, standard deviation) | 2 (1) | Assumption |
| RT-PCR sensitivity (average) | 71% | <sup>17</sup> |

Abbreviations: HR: Hazard Ratio; COPD: Chronic Obstructive Pulmonary Disease; ICU: Intensive Care Unit.

**eFigure 1.**
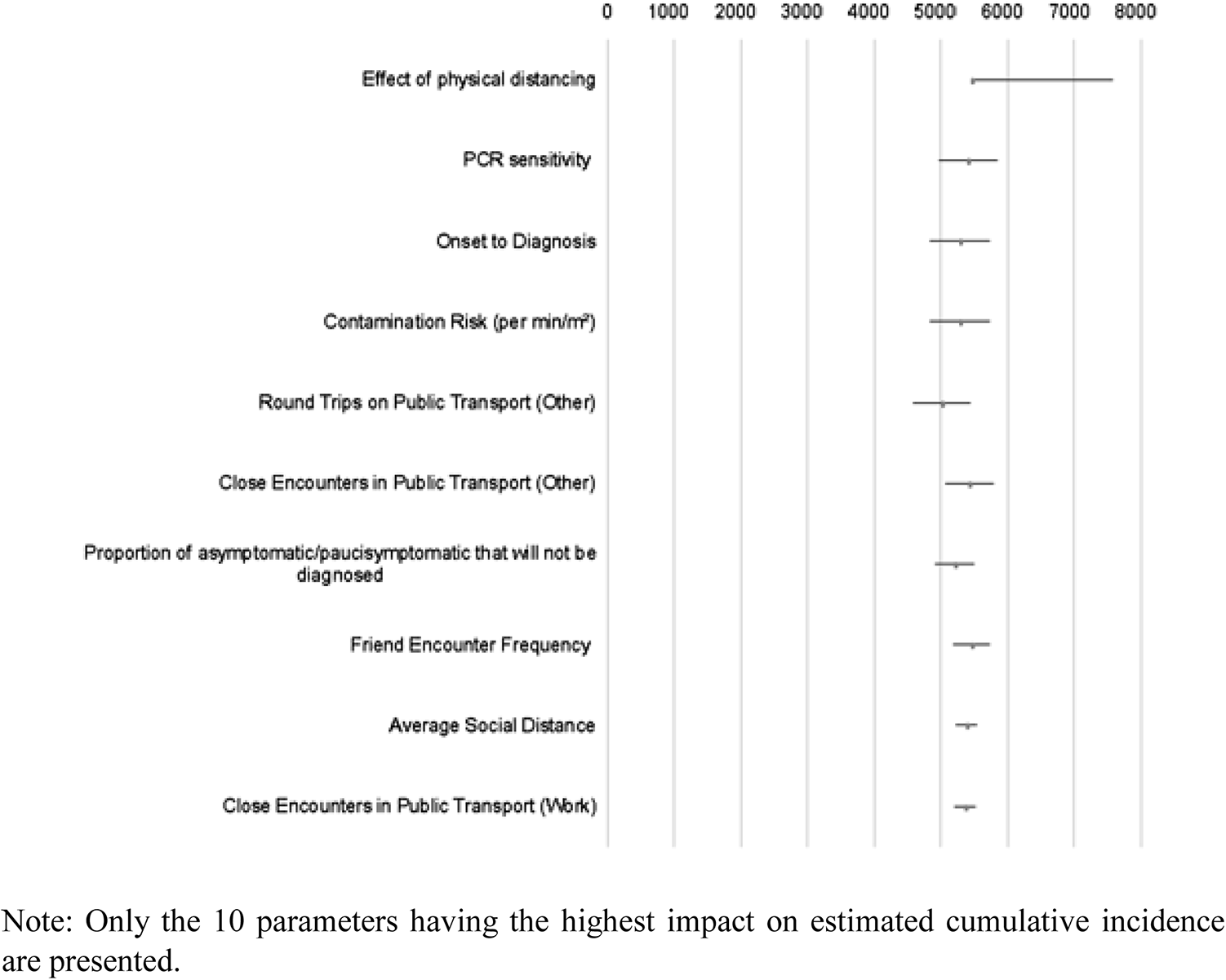
Sensitivity analysis: impact of varying by +/-20% each model parameter value on the estimated cumulative incidence for the combination of post-quarantine social distancing and mask-wearing for the general population, and shielding of vulnerable individuals.

**eFigure 2.**
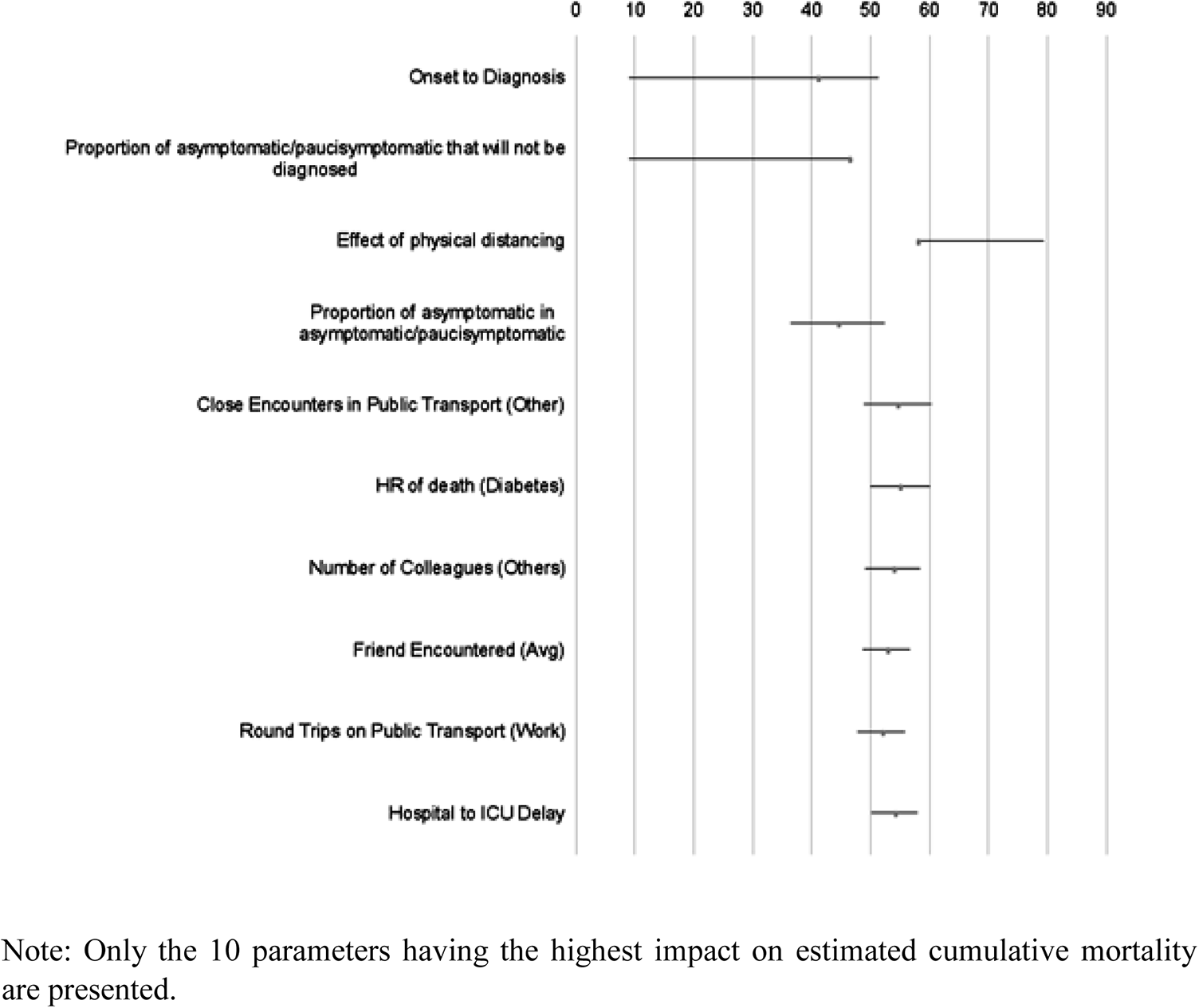
Sensitivity analysis: impact of varying by +/-20% each model parameter value on the estimated cumulative mortality for the combination of post-quarantine social distancing and mask-wearing for the general population, and shielding of vulnerable individuals.

**eFigure 3.**
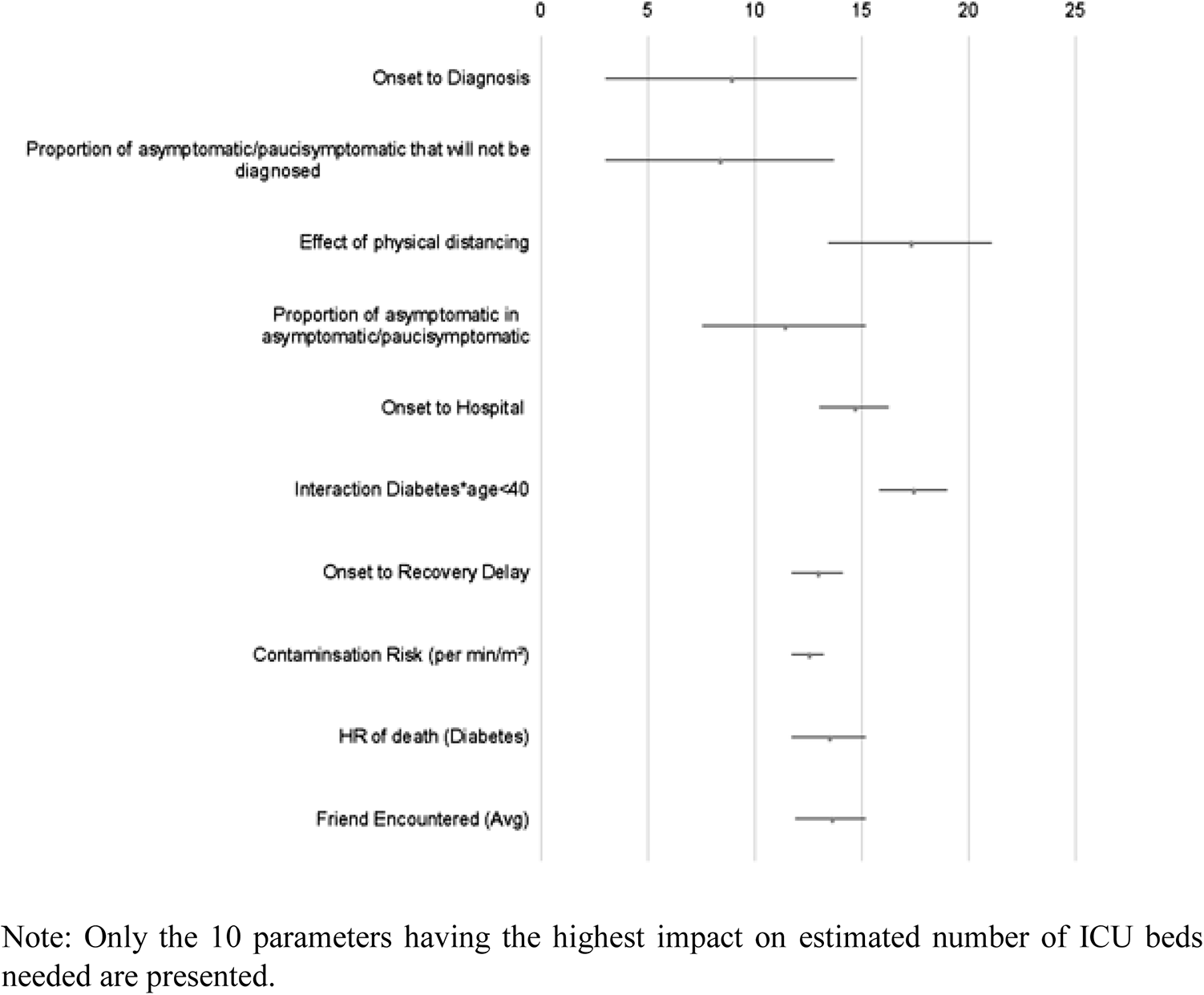
Sensitivity analysis: impact of varying by +/-20% each model parameter value on the estimated number of ICU beds needed for the combination of post-quarantine social distancing and mask-wearing for the general population, and shielding of vulnerable individuals.

## SUPPLEMENTAL TEXT SECTION

### Social contacts

Contacts were defined by their average duration (in minutes), their average distance (in meters), their frequency, and the number of individuals involved.^18–21^ For intrafamilial contacts, it was assumed that their average duration was 6 hours per day at a 1-meter distance every day for all household members. For contacts at school, outside the quarantine period during which these contacts were considered null, average duration was 6 hours at an average 2-meter distance, 5 days a week, for all classmates. Classmates were identified as children of the same age living in a similar location to represent the geographic clustering of schools. It was assumed that the average class size was 30. For contacts at work, outside the quarantine period during which these contacts were considered null, average contact duration with colleagues was assumed to be 7.5 hours at a 2-meter distance, 5 times a week. Only employed individuals aged 20 to 65 years had work-related contacts.^8^ We distinguished between small companies with 10 or fewer employees and regular or large ones.^8^ Individuals working in small companies had two colleagues on average, while employees of regular or large companies had an average of 10 colleagues. The number of colleagues was randomly drawn from a Poisson distribution. Work colleagues were identified at random within the city grid. For friends and family contacts, outside the quarantine period during which these contacts were considered null, it was assumed that the average duration was 180 minutes at a 1-meter distance, with one meeting a week on average. Outside the quarantine period, it was also considered that friend and family contacts occurred between households, for example, a couple with children could visit a friend’s or grandparent’s household.

Social networks were based on methods described by Gilbert et al.^10^ with a distance of 22 (Poisson distributed) in order to incorporate key aspects of social networks, such as the different sizes of personal networks, high clustering, positive assortment of degree of connectivity, and low density. Individuals were considered to visit the closest grocery store from their location 1.2 times a week, and meet an average of 5 people (Poisson distributed). Grocery stores were uniformly distributed throughout the city grid based on grocery stores’ density in France.^9^ Outside the quarantine period, contacts when going out of home were limited to cultural activities such as museum, sport, music or cinema events. It was assumed that contacts in restaurants or bars were captured through the friend and family contacts. The average number of times the family went out per year (Poisson distributed) was based on ticket sales’ from the French Ministry of Culture.^11^ Attendance at any public event was associated with a 120-minute duration at a 2-meter distance with an average of 5 individuals (Poisson distributed) randomly identified in the city grid. Finally, for public transport, we considered that all individuals used public transport 1.7 times a week for shopping or seeing family or friends. Workers were assumed to use public transport five times a week, twice a day (Poisson distributed). During public transport, a 30 min^22^ average duration at a 1-meter distance from a mean number of 3 to 5 individuals (Poisson distributed) randomly identified in the city grid was assumed.

Finally, it was also considered that the first patients were individuals infected via international travel. Thus, individuals could become infected through international contacts over time at a rate based on the frequency of infected patients that were initially diagnosed in France.^7^

### SARS-CoV-2 characteristics

A key uncertainty about COVID-19 is the proportion of infected individuals that are not diagnosed. Studies from China,^23^ Italy,^24^ and the United States^25^ suggest a high number of undiagnosed infections, ranging from 50% to 92% of all infections. Similarly, a study from France suggests that about 4 million people (representing about 6.0% of the French general population) were infected by the end of March 2020, contrasting with the 44,000 confirmed cases, suggesting a 1 in 100 diagnosis rate.^26^ This rate was confirmed by a recent study^12^ projecting 3.7 million (range: 2.3-6.7) people, i.e. 5.7% of the general population, will have been infected by 11 May. It was assumed that individuals with no or light symptoms (such as stomach pain or nausea) were not diagnosed, except if they were traceable contacts (i.e., intrafamilial, work, school) of diagnosed patients, and that all individuals with mild, severe, or critical symptoms were diagnosed. To reflect these assumptions, among infected individuals, the probability of being asymptomatic or lightly symptomatic in the model was set at 95% in children aged less than 10 years, since very few children have been diagnosed with COVID-19,^7^ and was assumed to decrease linearly with age. The slope of this decrease was calibrated to show a cumulative incidence (diagnosed + undiagnosed) of 1 in 100 diagnosis rate.

The probabilities of hospital admission (in case of severe symptoms), ICU admission (in case of critical symptoms) and death were based on estimates from Institut Pasteur.^12^ The probabilities of ICU admission and death were stratified by age and comorbidities, including hypertension, diabetes, obesity, coronary heart diseases, and chronic pulmonary diseases based on prior work.^2–6,12,14^ Delays between infection, symptom onset, hospital admission, ICU admission, death and recovery were based on prior reports^12,13,16,27^ and are detailed in **eTable 1**. Delays were randomly assigned based on the Weibull distribution.^28^

The risk of infection during a contact with an infected individual (per min/m^2^ of contact) was calibrated to reproduce the SARS-CoV-2 epidemiological data from France until the 22^th^ of April.^7^ A prior review^29^ suggested that the median of the basic reproduction number (R_0_) of COVID-19 would range between 1.40 and 6.49, with a median of 2.79, at the early stages of the epidemic. This variable was included as an outcome in our model to examine whether the predicted value was in line with published reports, and thus evaluate the potential predictive value of the model. The risk of transmission was assumed to be highest at the onset of symptoms and to decrease with time. To take into account the risk of transmission before developing symptoms,^16^ it was assumed that infected individuals were contagious starting one day after infection, albeit with a contagiousness that decreases exponentially the further away from onset. An exponential function was chosen because it fitted well with the dynamics of viral replication, based on these assumptions. Individuals who recovered were assumed to have acquired immunity against the virus and no longer at risk of infection. Based on prior work,^17^ sensitivity of reverse transcription polymerase chain reactions (RT-PCR) to detect COVID-19 cases was assumed to be 71%. We also assumed that the virus may not be sensitive to changes in temperature.

Finally, the number of ICU beds needed over time was compared to the number of ICU beds available in France, estimated at about 5,300 beds before the epidemic.^30^ However, following healthcare systems’ reorganization, the number of ICU beds has reached a total of 14,000 ICU beds in April.^30^ Because patients may require intensive care for other reasons than COVID-19 (e.g., stroke, myocardial infarction), we considered that excess ICU-bed occupancy corresponds to full occupancy of newly created ICU beds (i.e., 8,700 beds or 14 ICU beds per 100,000 people), and that the maximum number of ICU beds available for patients with critical symptoms of SARS-CoV-2 is 19 ICU beds per 100,000 inhabitant, assuming that they occupy 100% of newly created ICU beds and 75% of pre-existing ICU beds (i.e., 12,675 ICU beds). Patients requiring ICU level care with no available beds were assumed to have 100% probability of dying.

## Notes

### Competing Interest Statement

The authors have declared no competing interest.

### Funding Statement

This work did not receive any external funding.

